# Clinicians’ ratings of the Health of the Nation Outcome Scales (HoNOS) show sensitivity/bias that may affect patients’ progress: analyses of routine administrative data

**DOI:** 10.1101/2020.11.19.20234674

**Authors:** Jonathan Williams

## Abstract

**Objectives:** (1) To estimate clinician sensitivity/bias in rating the HoNOS. (2) To test if high or low clinician sensitivity determines slower resolution of patients’ problems or earlier inpatient admission.

**Design:** The primary analysis used many-facet Item Response Theory to construct a multi-level Graded Response Model that teased apart clinician sensitivity/bias from the severity of patients’ problems in routine HoNOS records. Secondary analyses then tested if patients’ outcomes depend on their clinicians’ sensitivity/bias.

**Outcome measures:** The outcome measures were (1) overall differences in sensitivity/bias between (a) individual clinicians and (b) different Community Mental Health Teams (CMHTs); (2) clinical outcomes, comprising (a) the rate of resolution of patients’ problems and (b) the dependence of the time to inpatient admission on clinician sensitivity/bias.

**Setting:** All archival electronic HoNOS records for all new referrals to all CMHTs providing mental health services in secondary care in a New Zealand District Health Board during 2007-2015.

**Participants:** The initial sample comprised 2170 adults of working age who received 5459 HoNOS assessments from 186 clinicians. From these initial data, I derived an opportunistic, connected, bipartite, longitudinal network, in which (i) every patient received HoNOS ratings from 2 or more clinicians and (ii) every clinician assessed more than 5 patients. The bipartite network comprised 88 clinicians and 778 patients; 112 patients underwent later inpatient admission.

**Results:** Sensitivity/bias differed importantly between individual clinicians and CMHTs. Patients whose clinicians had more extreme sensitivity/bias showed slower resolution of their problems and earlier inpatient admission.

**Conclusions:** Raw HoNOS ratings reflect the sensitivity/bias of clinicians almost as much as the severity of patients’ problems. Additionally, low or high clinician sensitivity can adversely affect patients’ outcomes. Hence, the HoNOS’s main value may be to measure clinician sensitivity. Accounting for clinician sensitivity could enable the HoNOS to fulfil its goal of improving mental health services.

**Strengths and limitations of the study:**

1. The study derived a connected network of clinicians and patients that approximates a rational design for estimating clinicians’ sensitivity/bias.
2. The opportunistic network sample was atypical, with chronic patients and experienced clinicians – so the study may *under*-estimate clinician bias.
3. The study’s statistical methods were appropriate to the ordinal nature of HoNOS ratings.
4. The study used earlier estimates of clinician sensitivity/bias to predict later outcomes – so that effects of clinician sensitivity/bias on outcomes may be causal
5. The study assumed that all HoNOS items tap a single dimension of the severity of patients’ problems.

## INTRODUCTION

The Health of the Nation Outcome Scales (HoNOS) aims to allow clinicians to summarise clinical problems in individual patients and to develop infrastructure for improving the quality of mental health care[1]. Collecting the HoNOS is now mandatory in some countries; in the UK, HoNOS ratings support ‘care clustering’ which may determine ‘Payment by Results’ (PbR – now the National Tariff Payment System).[2] However, the value of the HoNOS is controversial[3] and its psychometric and operational properties remain uncertain.[3–5] One possible reason for this controversy and uncertainty is variation (bias) between clinicians.[6] Here, I assessed clinician bias and its effect on the validity of HoNOS ratings, in routine use.

Initial field trials of the HoNOS reported good inter-rater reliability[1,7]. However, Ecob and colleagues later showed important clinician bias.[6] No subsequent studies have heeded their conclusion that “investigations of HoNOS scores *must* take assessors into account” (italics in the original).[6] Ecob’s analyses assumed that raw HoNOS scores are cardinal (e.g. that ‘Severe’ is exactly twice as bad as ‘Mild’), which may have distorted their results.[8,9] Here, I use mathematical methods that are appropriate to the ordinal nature of HoNOS ratings.[9]

The use of raw HoNOS ratings to inform PbR assumes that that not only individual clinicians but also teams in different localities use the HoNOS equivalently. However, there can be important differences between the HoNOS scores of different institutions or teams.[5,10–12] Hence, individual clinicians’ sensitivity/bias in rating the HoNOS may could reflect their team culture. I tested this by comparing clinician bias between Community Mental Health Teams (CMHTs).

A recent review found no evidence that the HoNOS improves services[3] and its predictive validity is uncertain.[13–15] Poor predictive validity may result if patient outcomes show non-linear dependence on clinician sensitivity/bias. In principle, *both* low *and* high clinician sensitivity/bias may cause adverse patient outcomes, because (a) insensitive clinicians (who give low ratings) may not recognise patients’ problems and so leave them untreated, but (b) over-sensitive clinicians (who give high ratings) may over-treat and so cause further problems.[16] I tested this by looking for quadratic (U-shaped) dependence of the rate of resolution of patients’ problems and of time to inpatient admission on estimates of their clinicians’ sensitivity/bias.

## METHODS

### Sample

The initial sample comprised all electronic HoNOS records from a District Health Board’s Mental Health Services for the period 02/07/2007 – 13/05/2015. All records were anonymised, archival, routine administrative data. So, there was no need to obtain prior consent to their use or prior ethical approval for the study. There was no public or patient involvement in the initial design or implementation of the HoNOS and since the present study is statistical, I did not seek such involvement, here.

Preliminary data cleaning eliminated duplicate records or assessments that rated six or fewer of the 12 HoNOS items. I then excluded ratings for patients (a) aged under 18 or over 65 at first contact, or (b) whose first HoNOS assessment was not a new referral to a CMHT, or (b) that occurred at discharge (including “discharge to another part of the service”) or at admission to the inpatient unit. (Supplementary Figure S1).

### Connected network sample

Estimating the biases of individual clinicians ideally requires a sample that comprises a connected network, with enough cross-linkage between clinicians and patients to allow information to flow freely.[17] Routine practice prevents cross-linkage, because clinicians usually rate the HoNOS only for patients whom they themselves assessed initially. However, clinicians sometimes rate each other’s patients, due to operational factors (e.g. if the primary clinician is on leave, etc.). I used this sporadic clinician-patient overlap to derive an opportunistic, connected bipartite network of patients and clinicians that had adequate cross-linkage (Supplementary Table S1 and Figure S2).

I estimated clinician bias using only HoNOS ratings from CMHTs. I selected only (i) patients who had at least two community HoNOS assessments by different CMHT clinicians; (ii) CMHT clinicians who rated at least 5 patients, of which (iii) they rated at least one patient in common with another clinician. I recursively eliminated patients and clinicians that failed criteria (i)-(iii), until a stable connected bipartite network formed. Finally, I added back to this sample any patients who received new referral or review ratings exclusively from the CMHT clinicians in the connected bipartite network (see Supplementary Figures S1-2).

### Admission sample

I assessed the validity of estimates of clinician bias and bias-adjusted estimates of patient severity by testing if they predict time to admission. Therefore, after defining and analysing the connected network to estimate clinician bias, I re-linked the patients in that sample with dates of inpatient admissions that occurred subsequently.

### Statistical analysis

All analyses used the open-source statistical programming language R [18]

### Estimation of clinician bias and severity of patients’ problems

I analysed the contributions of clinicians’ bias and the severity of patients’ problems to HoNOS ratings via Item Response Theory[17,19,20]. The analysis used a multi-level cumulative link model (mlclm). The mlclm’s fixed effects were patients’ demographic characteristics (age, gender, ethnicity), follow-up time and principal diagnosis (psychosis, mania, depression, anxiety, drug/ethanol abuse, personality disorder or dementia). The mlclm’s random effects estimated the severity of each patients’ problems and the sensitivity of each clinician and each item for detecting patients’ problems. The mlclm also included random effects for the items’ abilities to discriminate different degrees of the severity of patients’ problems. Hence, the mlclm was a multilevel graded response model (GRM)[20,21] (Supplementary Table S2) with a logit link that partitioned the raw HoNOS ratings (see Supplementary Methods and[20]) to reflect the following structure:-

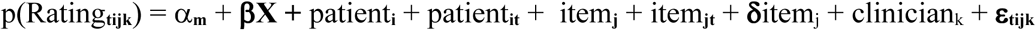

Where α_**m**_ are the ‘m’ thresholds between categories; **β** is the vector of coefficients for the **X** fixed effects, including follow-up time (which estimates differential test function for the demographic and diagnostic factors and follow-up); ‘t’ is follow-up time in each episode of care; patient_**i**_ is the random intercept for patient ‘i’ (the overall severity of the patient i’s problems at first assessment); patient_**it**_ is the random slope for patient ‘i’ over time ‘t’ (the rate of change of patient i’s problems over time); item_**j**_ is the random intercept for item ‘j’ (the mid-point of the item characteristic curve) at the first assessment; item_**jt**_ is the random slope of ratings on item ‘j’ over follow-up time (change in the likelihood of item j’s endorsement over follow-up ‘t’); **δ**item_j_ is the discrimination coefficient **δ** (the slope of the item characteristic curve) for each of the ‘j’ items[20,21]; clinician_**k**_ is the random intercept (overall bias/sensitivity) for clinician ‘k’; **ε**_**tijk**_ is error for each individual HoNOS rating. Estimation of the mlclm used Bayesian methods.[21,22] The mlclm estimated all the above effects simultaneously and so adjusted each effect for all of the others.

### Partitioning HoNOS ratings into clinician bias and severity of patients’ problems

I first tested if clinician bias determines mean HoNOS ratings and how well HoNOS ratings describe the severity of patients’ problems. These analyses used generalised additive modelling of location, shape and scale (gamlss), with the sinh-arcsinh distribution (see rationale, below). The first gamlss analysis regressed mean HoNOS ratings on the mlclm’s estimates of clinician sensitivity/bias; the second regressed the mlclm’s estimates of the initial severity of patients’ problems on mean initial HoNOS ratings. This analysis also computed the 2.5^th^ and 95^th^ centiles of the severity of problems for each category of HoNOS ratings.

### Comparison of bias between individual clinicians and CMHTs

I quantified clinician bias and variation between patients using the Median Odds Ratio (MOR).[23,24] I computed the bias for each CMHT as the mean of the mlclm’s random intercepts for each clinician in that CMHT. I also computed the mean severity of patients’ problems in each CMHT as the mean of mlclm’s random intercepts for patients receiving care from the CMHT. When clinicians’ or patients’ data contributed to more than one CMHT, I included them only in the CMHT to which they contributed most frequently. I combined the two smallest CMHTs into a single unit. I then compared the mean clinician sensitivity/bias and patient severities between CMHTs using Kruskal-Wallis (KW) χ^**2**^ tests.

### Assessment of predictive validity

I assessed the abilities of the mlclm’s estimates of the initial severity of each patient’s problems and of each clinicians’ sensitivity/bias (see above) to predict patients’ progress or time to inpatient admission (see Introduction). In particular, I tested if more extreme clinician sensitivity/bias predicted worse outcomes. So, I predicted that more extreme clinician sensitivity/bias would cause (a) more positive random slopes (i.e. slower resolution of patients’ problems), resulting in U-shaped dependence of random slopes on clinician bias; (b) earlier inpatient admission, resulting in *inverted*-U dependence of time to inpatient admission on bias. Therefore, I modelled the dependence of the outcomes on quadratic terms of clinician sensitivity/bias.

Analyses of change of the severity of problems used gamlss.[25] These models analysed all patients in the connected network, because the mlclm extracted random slopes of rates of change for all patients.

Analyses of time to inpatient admission estimated the survival of community status. [26] These analyses included only patients who later underwent inpatient admission, because including all patients in the analysis can introduce bias (by assuming that all patients will eventually become inpatients[26,27]). I estimated both accelerated failure time (AFT) and Cox’s proportional hazards (CPH) models.

Preliminary gamlss models that used the normal distribution fitted poorly – their residuals showed marked deviations from normality (both leptokurtosis and skewness). Such non-normality may occur in observational studies, as here, because unknown selection variables influence response distributions.[28] Empirically the sinh-arcsinh distribution (a flexible variant of the normal distribution, that can accommodate heavy-tailed combinations of mean, variance, skewness and kurtosis[29]) fitted the rate-of-change data well (Shapiro-Wilk normality test of residuals: W=0.999, p=0.9); the gamlss models used this distribution. AFT models used the Rayleigh distribution, since this may be optimal for distinguishing orthogonal components of a single measure (here, the separate roles of patients’ problems and clinician sensitivity in determining admission times).[30]

The gamlss analyses regressed both the mean and variance of the rates of change on the initial severity of each patient’s problems, clinician sensitivity, demographic and clinical variables. The survival models included only the mlclm’s estimates of the severity of patients’ problems and clinician sensitivity, because the sample was too small to include many covariates. The gamlss and survival analyses first tested if the initial severity of patients’ problems and clinician bias could determine the outcomes. They then tested if quadratic terms for clinician bias improved the model fits. I compared different models using Nagelkerke R^**2**^ and likelihood ratio χ^**2**^ (LR χ^**2**^) tests.

The mlclm’s estimates of clinician sensitivity/bias and patients’ problems may be statistically orthogonal (see Supplementary Figures S5a-b). However, there is a risk of causal circularity if later estimates of clinician sensitivity predict earlier patient outcomes. I minimised this risk in three ways: (1) I tested if the sensitivity of *only* the initial clinician determined the subsequent rate of change of patients’ problems. (2) Patients who underwent inpatient admission had earlier HoNOS assessments from 2 or more CMHT clinicians. So, I tested the dependence of time to admission on the mean sensitivity of all pre-admission clinicians. (3) Survival analyses excluded admissions which occurred less than 1 month after the first HoNOS assessment, in order to ensure that the initial HoNOS assessment preceded the decision to admit.

## RESULTS

### Sample

The clinicians in the connected network sample had rated more HoNOS records (median 27, IQR 17-52) than their unselected colleagues (median 16, IQR 3-45). The patients in the selected sample differed from the rest in three ways that may reflect the selection process: the selected patients received (1) more HoNOS assessments, (2) from more clinicians, (3) over a longer period of follow-up (Table 1). The connected network sample also showed differences that did not relate to the selection process: its patients included lower a proportion of males and a higher proportion of of non-Europeans, compared with the unselected patients (Table 1).

**Table 1:** the clinical characteristics of the unselected, connected and admission samples.

| Clinical measure | Sample: | unselected | Connected-mlclm | Admission sample |
| --- | --- | --- | --- | --- |
| Age at 1<sup>st</sup> contact |  | 37 (26-48) | 38 (27-48) | 35 (25-47) |
| Gender (Female : Male) |  | 1142 : 1028 | 449 : 329<sup>a</sup> | 46 : 66<sup>b</sup> |
| Ethnicity (European : other) |  | 1108 : 1062 | 262 : 516<sup>a</sup> | 50 : 62<sup>b</sup> |
| Setting (CMHT : IPU : both) |  | 1420 : 78 : 672 | 653 : 0 : 125<sup>1</sup> | 0 : 0 : 112 |
| Number of assessments |  | 2 (1-7) | 4 (3-8)<sup>a</sup> | 13 (9-18)<sup>b</sup> |
| Follow-up duration (years) |  | 0.1 (0-2.2) | 1.3 (0.5-3.4)<sup>a</sup> | 2.18 (1.08-2.92)<sup>b</sup> |
| Total HoNOS scores |  | 0.7 (0.4-1.1) | 0.7 (0.4-2.0) | 0.80 (0.63-1.02)<sup>b</sup> |
| Number of HoNOS raters |  | 1 (1-3) | 3 (2-4)<sup>a</sup> | 6 (4-8)<sup>b</sup> |

Values for age, number of assessments, follow-up, total HoNOS scores and number of HoNOS raters are medians and inter-quartile ranges (IQRs); values for gender, ethnicity and setting are counts. Key: a – differs from non-selected sample; b – differs from connected network sample; 1 – note that the mlclm analysis of the connected network sample used only HoNOS ratings that patients received in community settings. Abbreviations: CMHT = Community Mental Health Team; IPU = Inpatient Unit; mlclm multi-level cumulative link model.

### Partitioning raw HoNOS ratings into patient problems and clinician bias

The mlclm’s predicted scores – including its random effects – fitted the progress of individual patients’ mean HoNOS ratings adequately (Supplementary Figures S3-4).

Clinician bias accounted for two-fifths of the variance in mean HoNOS ratings both at initial assessment (38% – Figure 1a) and during follow-up (40%). The mlclm’s estimates of initial severity of patient problems accounted for half of the variance of mean HoNOS ratings at initial assessment (51%) and during follow-up (56%) (Supplementary Figure S5a; see Figure 1b). So, together, the mlclm’s estimates of clinician sensitivity and the severity of patients’ problems accounted for almost all the variance in mean HoNOS ratings (89% initially; 96% during follow-up).

**Figure 1:**
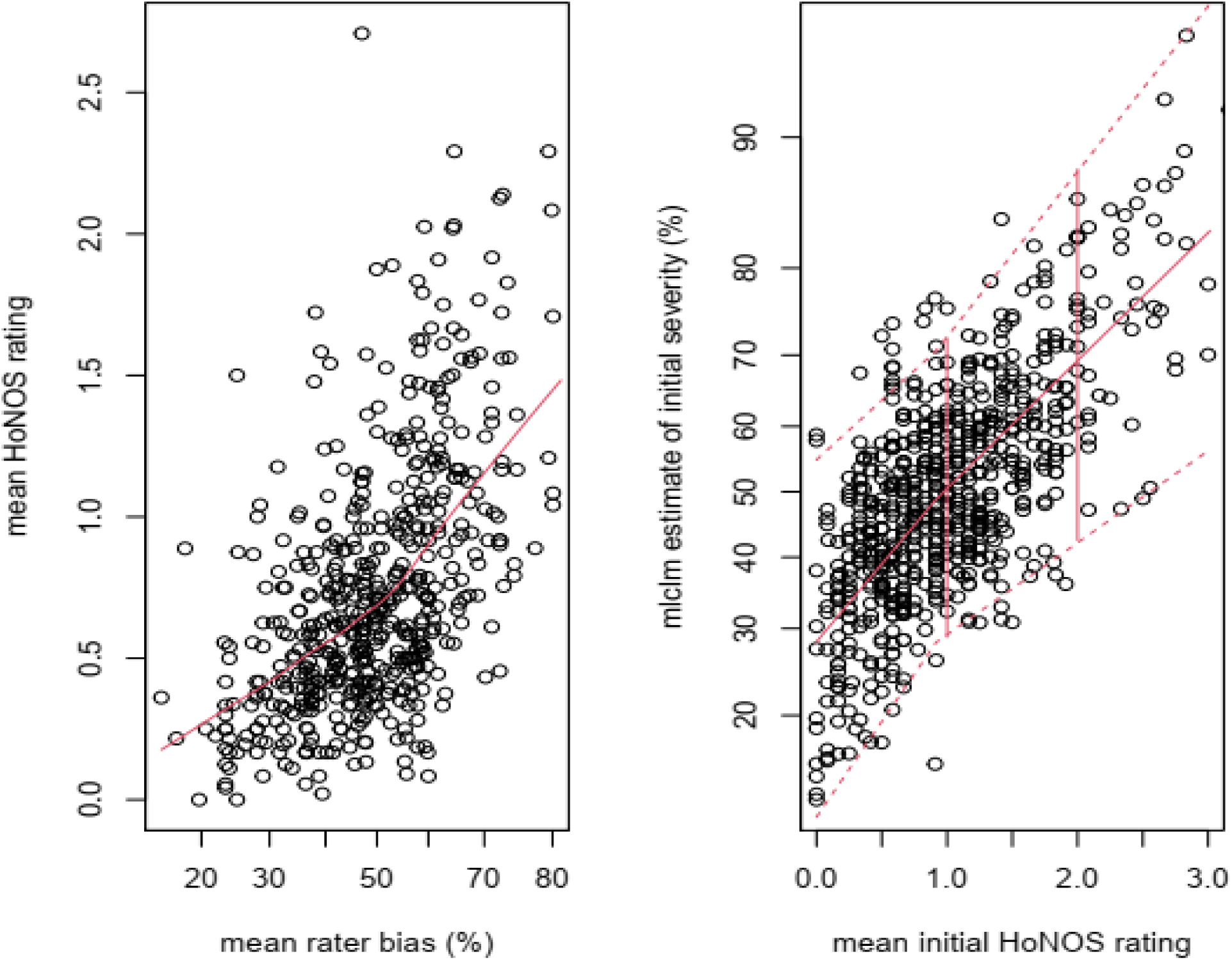
Relations between mean HoNOS ratings and mlclm random effects. Legend: Figure 1a shows the dependence of mean raw HoNOS ratings (y-axis) on clinician sensitivity/bias (x-axis). More sensitive clinicians give higher HoNOS ratings. The values of sensitivity are percentages of the total possible range (0% for a clinician who always rates every HoNOS item as “none”=0; 100% for a clinician who always rates every item as “severe”=4). The red line shows the smoothed median raw HoNOS rating at each level of clinician sensitivity/bias. Figure 1b shows the dependence of the mlclm’s estimates of the initial severity of patients’ problems (y-axis) on mean raw initial HoNOS ratings (x-axis). The values for severity are percentages of the total possible range (corresponding to patients who always receive HoNOS ratings of either none=zero or severe=4, for all 12 scales). The dashed red lines show the 2.5th and 95^th^ centiles of initial severity at each mean raw HoNOS rating. The vertical red lines show the 95% Confidence Limits of the ranges of initial severity that correspond to mean raw initial HoNOS ratings of 1=questionable or 2=mild).

Initial HoNOS ratings accounted for about half the variance of the mlclm’s estimates of the initial severity of patients’ problems (47%; Figure 1b). In concrete terms, (a) in any given HoNOS category, the 95%CI of estimates of severity spans almost half their range (see Figure 1b); (b) a patient who receives ‘none’ ratings for all HoNOS items from an insensitive clinician and a second patient who receives ‘mild’ - to - ‘moderate’ ratings for all items from an over-sensitive clinician may both have average (50%) bias-adjusted levels of severity of their problems (see further description of clinician sensitivity/bias, below).

Even though the mlclm’s estimates of clinician sensitivity/bias and of the severity of patients’ problems related strongly to total HoNOS scores, the estimates for clinicians and patients were effectively orthogonal (both Spearman’s **ρ** < 0.05, both p > 0.3). (See Supplementary Figures S6a-b).

### Reliability

#### Differences between individual clinicians

There was important variation between clinicians (median odds ratio, MOR = 2.0, 95% Credible Interval = 1.6-2.6; Figure 2). In concrete terms, (a) on average for random pairs of clinicians, if the less sensitive rates 50% of 100 patients’ problems as ‘mild’ and 50% as ‘moderate’ (1:1), then the more sensitive would rate 33% of the *same* patients’ problems as ‘mild’ and 67% as ‘moderate’ (2:1); (b) the 95% CIs of lowest and highest tertiles of clinician sensitivity/bias did not overlap (Fig. 2); (c) the variation between clinicians was similar to that between patients at their initial assessment (MOR = 2.2, 95% CI = 1.7-2.8) (but it was less than differences between patients’ rates of change (MOR = 2.7, 95% CI = 2.0-3.8) (Supplementary Figs S7a-b).

**Figure 2:**
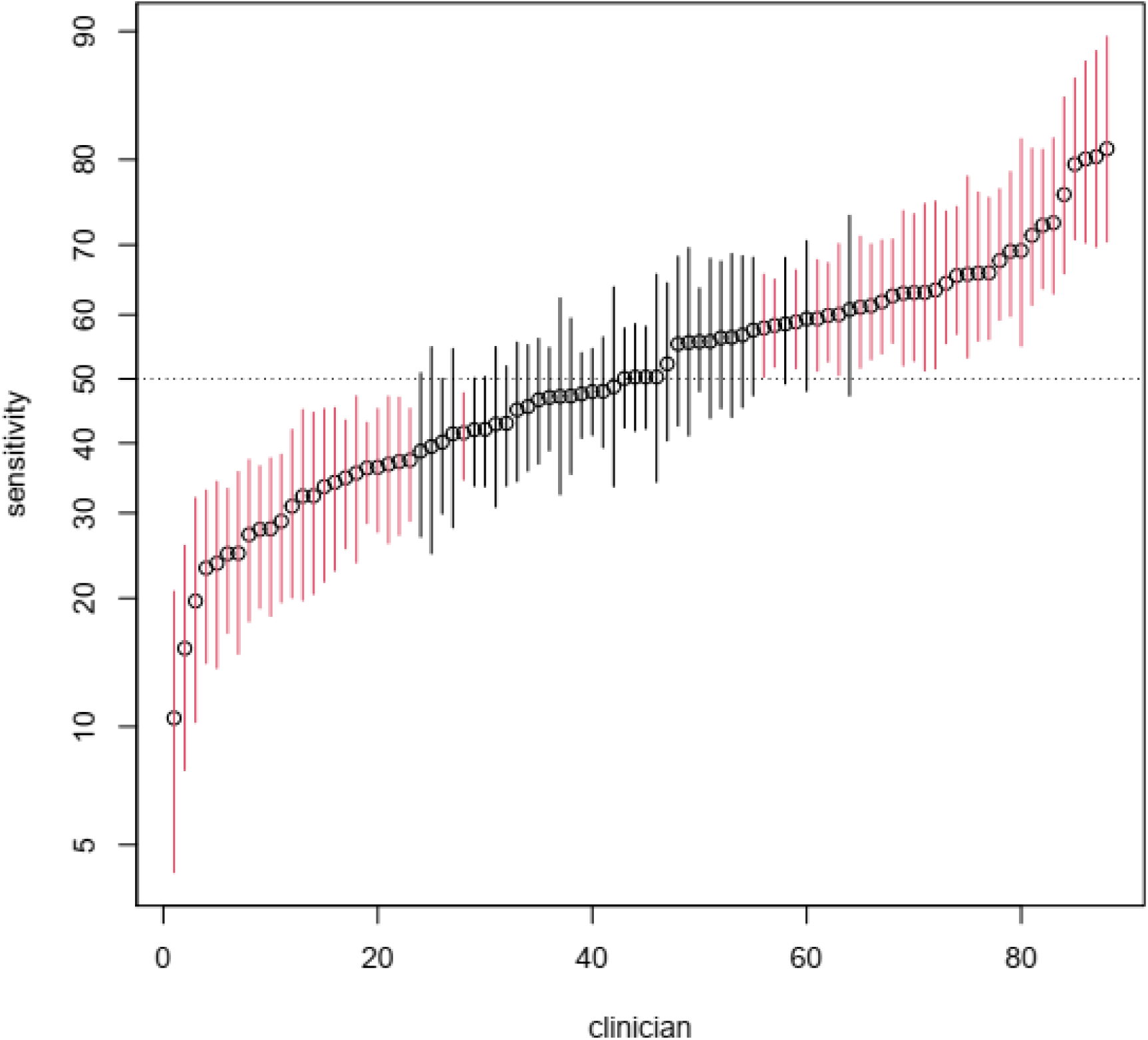
The sensitivities (bias) of individual clinicians. Legend: The caterpillar plot’s y-axis shows the estimate of sensitivity for each clinician, together with its 95% Credible Interval from the Bayesian multi-level cumulative link model (mlclm). The scale of the y-axis is percentage sensitivity when rating HoNOS items (see details in legend of Figure 1). The x-axis shows the index for each clinician, ordered by increasing sensitivity. CIs in red do not overlap the average value (50%); less than one third of raters are ‘average’.

#### Differences between Community Mental Health Teams

Clinician bias differed substantially, but not quite significantly, between CMHTs (KW χ^2^ = 7.0, 3df, p=0.07; Figure 3). After omitting a single outlier with very low sensitivity from Team 3 (see Figure 3), these differences were significant (KW χ^2^ = 8.9, 3df, p=0.03). The initial severity of patients’ problems and their annual rates of change were both similar between CMHTs (initial severity: KW χ^2^ = 0.3, 3df, p=0.96; annual change: KW χ^2^ = 4.4, 3df, p=0.22; Supplementary Figures S8a-b).

**Figure 3:**
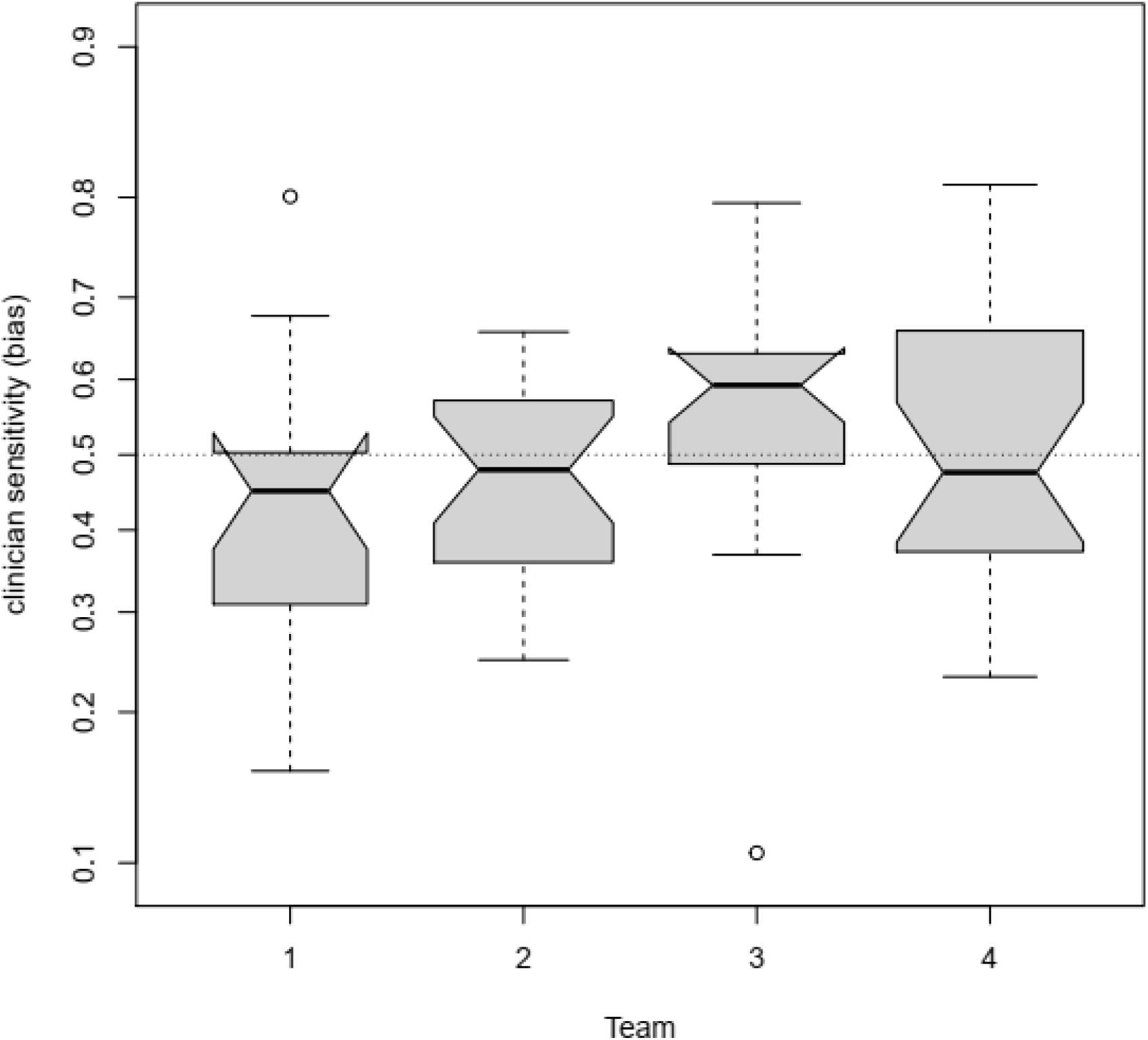
Estimates of clinician sensitivity, grouped by CMHTs. Legend: The notched box-and-whisker plots show the median and inter-quartile ranges of estimates of the clinician sensitivity (the random intercepts for clinicians in the mlclm). The whiskers cover 99% of the range predicted from the inter-quartile range. The notches represent the confidence intervals for the medians, so that two CMHTs whose notches do not overlap are likely to be differ significantly. The widths of the boxes are proportional to the numbers of each CMHT’s clinicians.

### Predictive validity

#### Rate of change of severity of problems

Patients whose initial clinicians had more extreme sensitivity/bias showed slower resolution of their problems (Figure 4). Statistically, the rate of reduction of patients’ problems showed U-shaped dependence on clinician bias (gamlss: LR χ^2^=4.31, 1df, p=0.038, 95%CI = 0.004 – 0.126; Figure 4). Also, patients with more severe initial problems improved faster (i.e. the mlclm’s random slopes related inversely to its random intercepts: t = −9.7, 744df, p<0.001; Supplementary Figure S8).

**Figure 4:**
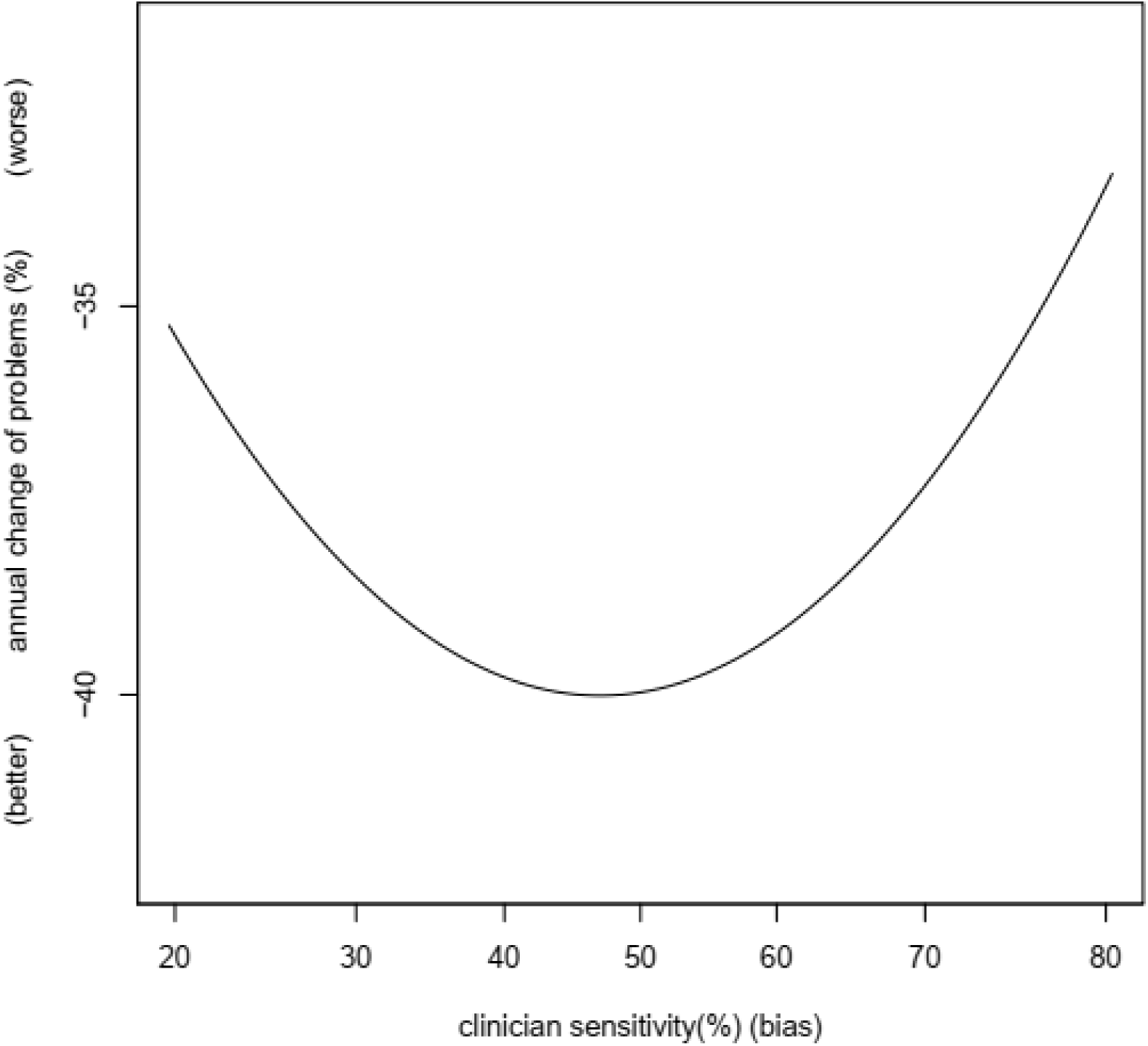
Dependence of annual change of severity of patients’ problems on clinician sensitivity. Legend: The x-axis represents the sensitivity of the first clinician to rate each patient (random intercepts for clinicians, from the mlclm). The graph truncates the range of clinician sensitivities at 1% and 99% of the full observed range. The y-axis represents the annual change of severity of patients’ problems (random slopes across time for patients, from the mlclm). More negative slopes (smaller y-values) indicate faster resolution of patients’ problems (better outcomes).

#### Time to admission

Among patients who underwent inpatient admission, the first admission occurred earlier in those whose pre-admission CMHT clinicians had more extreme sensitivity (Figure 5) (AFT: LR χ^2^ = 11.5, 1df, p=0.0007, 95%CI = −0.707 – −0.213; CPH: LR χ^2^ = 5.1, 1df, p=0.024; see also Supplementary Figure S10).

**Figure 5:**
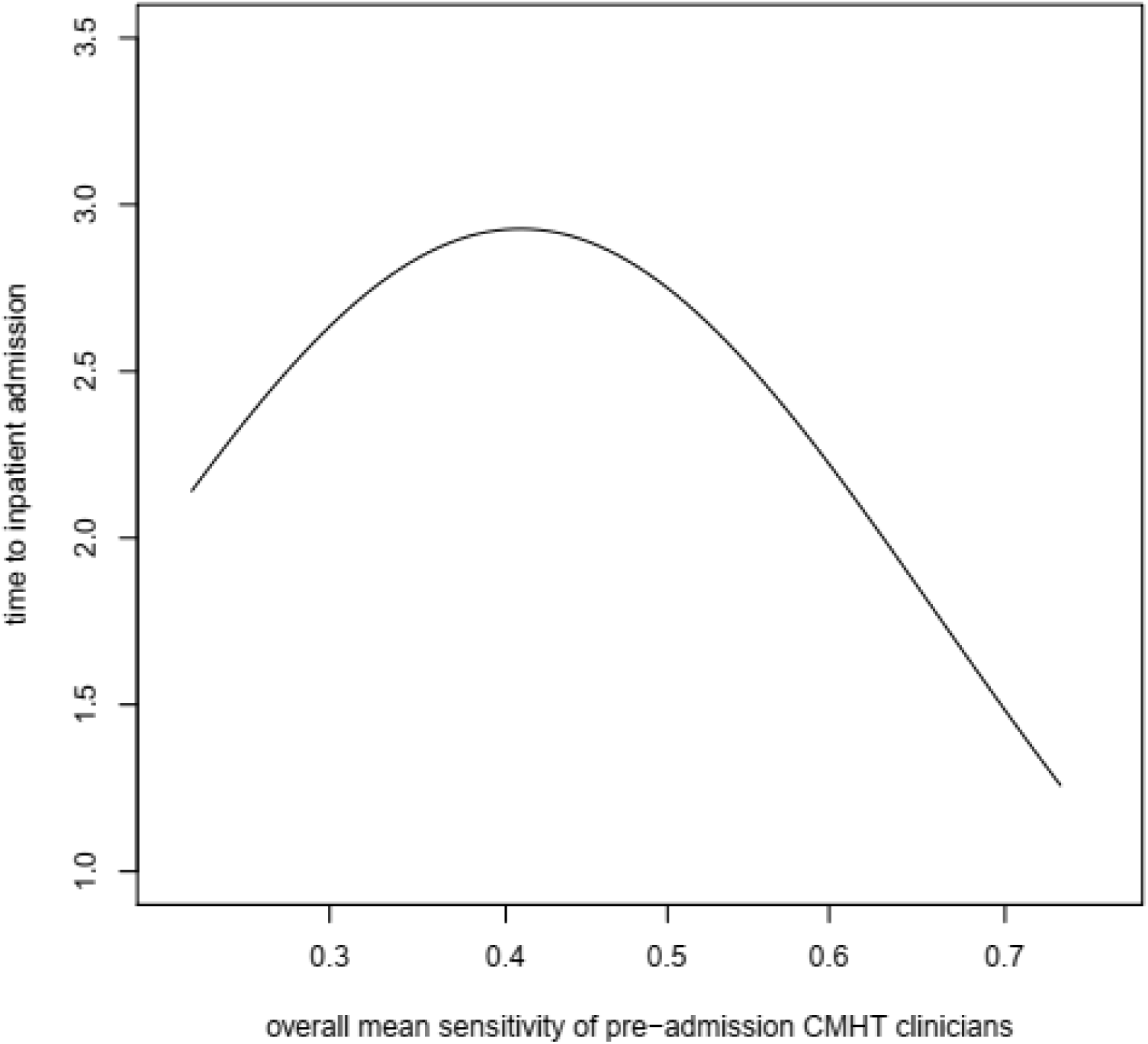
Dependence of time to inpatient admission on clinician sensitivity. Legend: The y-axis represents the average duration of community status for the group of patients who underwent inpatient admission. The x-axis represents the mean sensitivity of all CMHT clinicians who rated each patient before inpatient admission. The graph truncates the range of clinician sensitivities at 1% and 99% of the full observed range. Patients whose CMHT clinicians, overall, had the lowest or highest levels of sensitivity/bias had shorter intervals to inpatient admission (worse outcomes).

## DISCUSSION

### Principal findings

Clinician bias importantly influences raw HoNOS scores in routine practice. In fact, (a) raw HoNOS scores provide almost as much insight into clinicians’ sensitivity to their patients’ problems as into the actual severity of those problems; (b) any given raw HoNOS rating can associate with almost half the range of the bias-adjusted severity of patients’ problems. Importantly, this clinician sensitivity/bias may affect patient progress and management. In particular, patients whose clinicians showed more extreme bias (low, or high sensitivity) had worse outcomes, with slower resolution of their problems and earlier inpatient admission.

### Strengths and weaknesses

The study’s main strength is that it derived a connected network of patients and clinicians that approximated the rational design for assessment of clinician bias. Additionally, (1) the mlclm accounted for the ordinal nature of HoNOS ratings and so should outperform the linear analysis of the only comparable study.[6,8,9] (2) The mlclm relaxed the assumption (implicit in summing raw HoNOS ratings) that all HoNOS items contribute equally to estimating the severity of patient’s problems. (3) The mlclm provided independent estimates of the severity of patients’ problems and of clinician bias, that were effectively orthogonal. (4) The study used earlier clinician bias to predict later outcomes, so effects of bias on outcomes could be causal (but see below). (5) The study included an important objective outcome – time to admission.

The study’s main weakness is that both the patients and clinicians in the connected network were atypical: the patients were more chronic and the clinicians had completed more HoNOS assessments than their unselected peers. However, these atypical features should *reduce* any bias in clinicians’ HoNOS ratings. So, the study may *under-*estimate clinician bias and its effects in routine practice. Further weaknesses are: (1) The mlclm assumes that all HoNOS items tap a single underlying latent dimension of the “severity of patients’ problems”. This assumption conflicts with reports that the HoNOS has a multi-dimensional structure.[31,32] However, those findings are themselves inconsistent (e.g. compare [33–35]) and a bifactor analysis indicated that “HoNOS can be used as a unidimensional scale”.[33] Further studies should test if the quadratic dependence of patient outcomes on clinician bias depends on this simplifying assumption. (2) Clinician sensitivity/bias in rating the HoNOS is only important if it reflects clinician factors that *cause* patient outcomes. [16,36,37] The fact that the study’s measures of clinician sensitivity/bias preceded changes in outcomes allows the possibility of causality. However, the causal mechanisms are unclear, because most clinicians were Community Mental Health Nurses (CMHNs), who did not make diagnoses, prescribe medication or hold admission rights. Even so, CMHNs’ opinions are often crucial in determining inpatient admissions and clinician sensitivity here strongly influenced time to inpatient admission. This supports the possibility that clinician sensitivity/bias can cause outcomes, but further work should test this possibility more rigorously.

The graded rating scale model (GRM) that I used here assumes, like total HoNOS scores, that there is a single dimension of the severity of patients’ problems;[20] but GRM relaxes the additional assumption of total HoNOS scores that each item contributes identically to estimating overall severity. Ideally, the analysis should allow ‘differential item function’ (DIF) in relation to diagnostic category, because some HoNOS items tap the principal symptoms of some diagnostic categories (e.g. psychosis, depression and problems with ethanol or drugs). However, the numbers of patients in diagnostic categories in the present opportunistic sample may be too small to support accurate estimation of DIF (see[20]). Larger studies should adjust for DIF in HoNOS items and perhaps also in rater characteristics (e.g. experience).

### Comparison with previous reports

The present finding of important variation between clinicians converges with that of Ecob and colleagues[6] and with general evidence that raters often show important bias[17,38–40]. My results strongly reaffirm Ecob’s caveat that users of HoNOS ratings “*must*” take assessors into account.[6] Here, differences between individual clinicians almost equal those between the severities of individual patients’ problems, indicating that clinicians use the HoNOS in different ways.[41–43] In effect, raw HoNOS ratings measure clinicians’ opinions as much as their patients’ problems.

The present study bears little comparison with previous reports that did not adjust for clinician sensitivity/bias, because this adjustment extracts so much variance from raw HoNOS ratings. The present results indicate that only half the variance of raw HoNOS scores reflects the target signal – the severity of patient problems. Hence, the signal-to-noise ratios for analyses of raw HoNOS ratings in all previous reports may be approximately 1:1. Such low signal-to-noise may explain many discrepancies between those reports’ results (e.g. see Table 4 in[33]). However, adjustment for clinician sensitivity/bias, as here, may reduce unexplained ‘noise’ to 5-10% of HoNOS scores’ total variance, and so increase signal-to-noise ratios for estimates of patients’ problems to 5:1 – 10:1. Such strong signal should support much more robust reliability of HoNOS ratings, in many settings (see further, below).

Different CMHTs showed different levels of clinician bias. This is consistent with previous reports of variations between hospital and CMHT clinicians,[5] and at a broader level between hospitals and general practices.[11,44] Together, these results indicate that routine HoNOS ratings can detect important cultural differences in sensitivity/bias between providers – which may be helpful for planners and funders (see further, below).

The present findings that more extreme clinician sensitivity/bias related to worse patient outcomes are novel. Edbrooke-Childs and colleagues reported substantial variation in rates of treatment drop-out between individual clinicians,[41] but they did not relate this variation to the individual clinicians’ biases. Previous proposals that more extreme clinician sensitivity may determine adverse patient outcomes were qualitative.[16,36,45] The present findings provide a means to quantify the importance of these *a priori* principles.

### Implications for clinicians and planners

#### Clinicians

The present results indicate that raw HoNOS ratings are unhelpful, clinically. However, since clinician sensitivity/bias accounts for almost half of the variance of raw HoNOS ratings, adjustment for this sensitivity could allow accurate estimation of the true severity of patients’ problems. Bias-adjusted HoNOS scores could even be accurate enough to provide meaningful insights when providing treatment and prognosis for individual patients (cf [46]).

#### Planners

The present and previous evidence[6] that clinician sensitivity/bias contaminates raw HoNOS ratings has major implications for planners. Raw HoNOS ratings are the basis of allocating cases in “care clustering”, which determines “Payment by Results” (PbR) in the UK. There is already concern that care clusters are inadequate to support PbR[47] and clinician bias may further impair care clustering. Although there have been calls for training to standardise the use of HoNOS,[48] there is little evidence that training is effective[38,49–52] (and reducing clinicians’ bias/sensitivity could reduce the utility of their HoNOS ratings – see below). Overall, therefore, the present results militate against using *raw* HoNOS ratings to inform PbR. In contrast, bias-adjusted HoNOS ratings may reliable enough to support PbR. Additionally, adjustment for bias at the levels of both individual clinicians and whole teams would preclude possibilities that either cultural differences between providers or “gaming” could attract disproportionate funding.[53] Therefore, adjusting PbR to account for clinician sensitivity/bias may improve efficiency in mental health services.

### Further Research

(1) The present study establishes the principle that clinician sensitivity/bias may affect patient progress. However, the sample was unrepresentative and too small to establish levels of clinician sensitivity that yield optimal patient outcomes. Larger studies could address this crucial issue, by capitalising on natural variation in sensitivity between clinicians. (2) The present study analysed only effects of patient factors on HoNOS ratings. The present methods can readily generalise to include factors relating to service provision (e.g. type of treatment, profession of clinician), social settings (e.g. neighbourhood deprivation indices) and financial or legal outcomes (e.g. benefit use, criminal justice contacts). (3) Adjusting HoNOS scores for clinician bias should greatly improve the accuracy of estimating the severity of patients’ problems. Hence, analyses of bias-adjusted routine HoNOS ratings could assess therapeutic interventions in large-scale real-world practice, rather than restricting trials to small, unrepresentative, highly-selected samples[54,55]. Finally, (4), disentangling the contributions to HoNOS scores of the severity of patients’ problems and clinicians’ biases could importantly improve clinical governance and leadership. Specifically, the present methods could help to develop (a) “control limits”,[56] to avoid unnecessary attention to random negative variation in provider performance;[56] (b) positive leadership[57], by defining and disseminating aspects of clinicians’ routine practice that improve patient outcomes. In summary, re-designing services to assess and account for clinician bias could finally justify the costs of routine HoNOS collection (cf [58]).

## Supporting information

all supplementary information

## Data Availability

Data will be freely available from the internet

https://doi.org/10.7910/DVN/NSSZT2

## Author roles

JHW was responsible for all aspects of the study: the concept, all data analysis and drafting the manuscript

## Data sharing statement

The anonymized data, code to generate the statistical models and the models themselves are available at https://doi.org/10.7910/DVN/NSSZT2

## Acknowledgements

I am grateful to Dr Murray Patton for allowing access to the data.

## Registration

I did not pre-register the protocol for these analyses. However, they follow from previous work that I presented at the Royal Australia and New Zealand College of Psychiatrists Annual Congress and I outlined the idea for the present work to Dr Arran Culver at the New Zealand Ministry of Health in Wellington in February 2020.

