## Supplementary material for "Clinicians’ ratings of the Health of the Nation Outcome Scales (HoNOS) show sensitivity/bias that may affect patients’ progress: analyses of routine administrative data": all supplementary information: BMJO Supplementary tables figures and legends 191120.pdf

**Table S1**: measures of linkage in the bipartite network sample, for each CMHT

| Community Mental Health Team | Connectance (%) | Chequerboardness (%) |
|---|---|---|
| CMHT 1 | 4.8 | 86.3 – 92.2 |
| CMHT 2 | 4.3 | 87.6 – 90.0 |
| CMHT 3 | 3.6 | 87.5 – 96.5 |
| CMHT 4 | 2.8 | 93.4 – 96.8 |

Legend: The 4 CMHTs are those which appear in the Main Report (see Figure 2). Connectance is the proportion of possible links in the bipartite network that actually exist and reflects clinician-patient overlap (if every clinician rated every patient, then connectance would be 1). Chequerboardness averages the number of instance of exclusive (01/10) links for all pairwise clinician-patient combinations and so reflects continuity of care (if each patient sees only one clinician, then chequerboardness would be 1). Computation of these statistics used the R package 'bipartite' [1,2]


1 Dormann CF, Fründ J, Blüthgen N, *et al.* Indices, Graphs and Null Models: Analyzing Bipartite Ecological Networks. *The Open Ecology Journal* 2009; **2**. https://benthamopen.com/ABSTRACT/TOECOLJ-2-1-7.

2 Dormann C. Using bipartite to describe and plot two-mode networks in R. 2020. https://cran.r-project.org/web/packages/bipartite/vignettes/Intro2bipartite.pdf


**Table S2**: differences between HoNOS items in estimating overall severity of patients' problems

| Item | Measure: | Intercept | Discrimination | Slope over time |
|---|---|---|---|---|
| Agitation/aggression | | -0.04 | 0.03 | -0.15 |
| Self-harm | | -0.35 | -0.13 | -0.38 |
| Drug/alcohol problems | | -0.51 | -0.38 | -0.00 |
| Cognitive impairment | | -0.31 | 0.13 | 0.11 |
| Physical illness | | -0.40 | -0.25 | 0.14 |
| Psychotic symptoms | | -1.36 | -0.25 | 0.32 |
| Depressive symptoms | | 1.20 | 0.06 | -0.28 |
| Other symptoms | | 1.42 | -0.06 | -0.16 |
| Family problems | | 0.49 | 0.26 | 0.08 |
| Problems with ADLs | | 0.12 | 0.21 | -0.00 |
| Accommodation problems | | -0.43 | 0.15 | 0.12 |
| Problems with activities | | -0.03 | 0.14 | 0.13 |

**Legend**: the values in the table are deviations from the overall mean. The Intercepts represent the differences between the contributions of HoNOS items to the overall severity of each patient's problems at first assessment (when follow-up time=0, within each episode of care). The slopes over time are rates of change of the contribution of each HoNOS item to the severity of each patient's problems (for example, the problems of chronic patients may reflect psychotic symptoms more than depressive symptoms). Discrimination coefficients reflect each item's ability to gauge different levels of severity

**Figure S1**: the selection of the sample for analysis

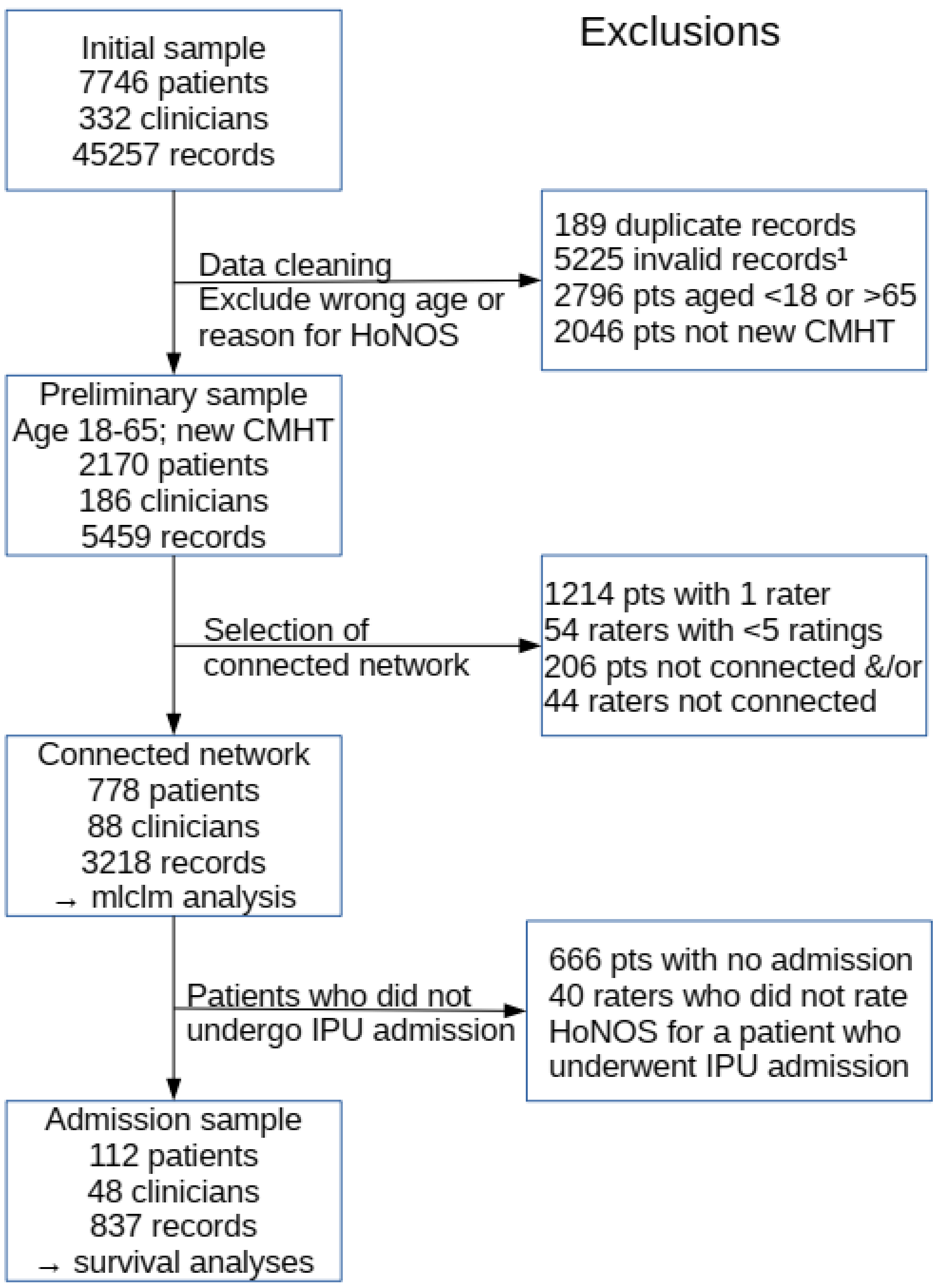


**Legend**: The diagram shows the numbers of HoNOS records, patients and clinicians (raters) at each stage of the selection process. Note that the table may show exclusion of the same record twice – for example, a record excluded from the connected network because it was invalid and/or rated a patient aged over 65. Notes: 1 – duplicate HoNOS records were for the same patient, at the same time, for the same HoNOS reason in the same setting; 2 - invalid HoNOS records were those with ratings outside the range 0-4 (none=0, questionable=1, mild=2, moderate=4, severe=4) or with more than 6 missing ratings. Abbreviations: pts = patients; raters = clinicians; CMHT = Community Mental Health Team; IPU = Inpatient Unit.

**Figure S2**: an example connected bipartite network

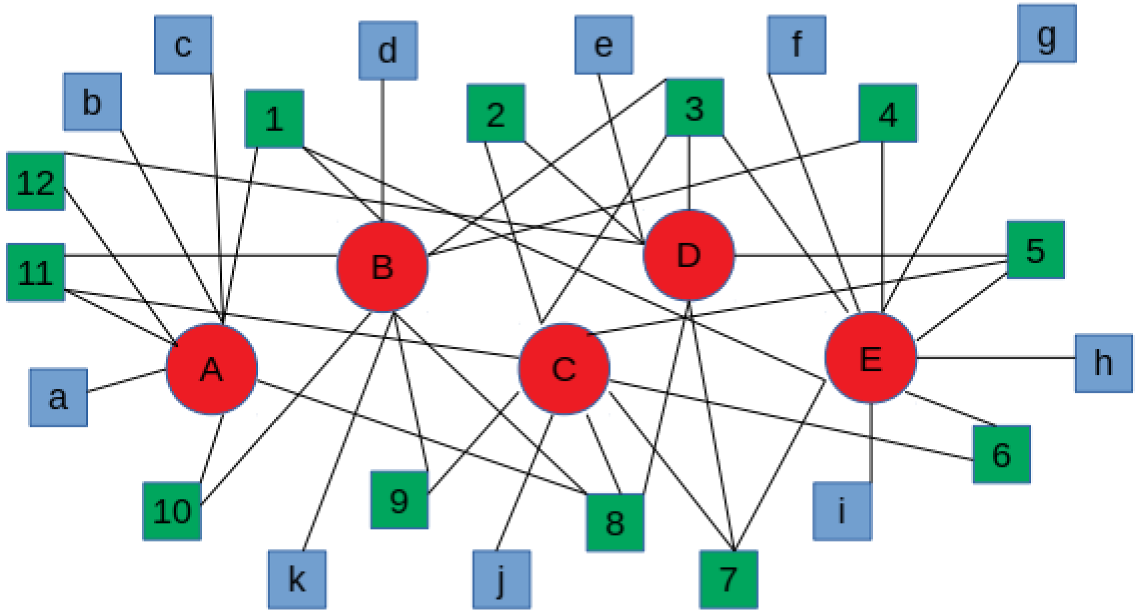


**Legend**: The figure shows an example bipartite network analogous to the sample for the multi-level cumulative logit model. I initially derived a connected network in which every (a) clinician rated at least 5 patients, (b) every patient received ratings from at least two clinicians and (c) every clinician rated at least one patient in common with at least one other clinician. I then added back to the connected network any patients who received HoNOS ratings exclusively from clinicians in the network. In the figure, the red circles (A-E) represent clinicians; green circles (1-12) represent patients who received HoNOS ratings from more than one clinician (the connected bipartite network); blue circles (a-k) represent patients who received HoNOS ratings from only one clinician, who was part of the connected network; links between nodes represent HoNOS assessments.

**Figure S3**: Comparison between actual and predicted mean HoNOS scores for individual patients

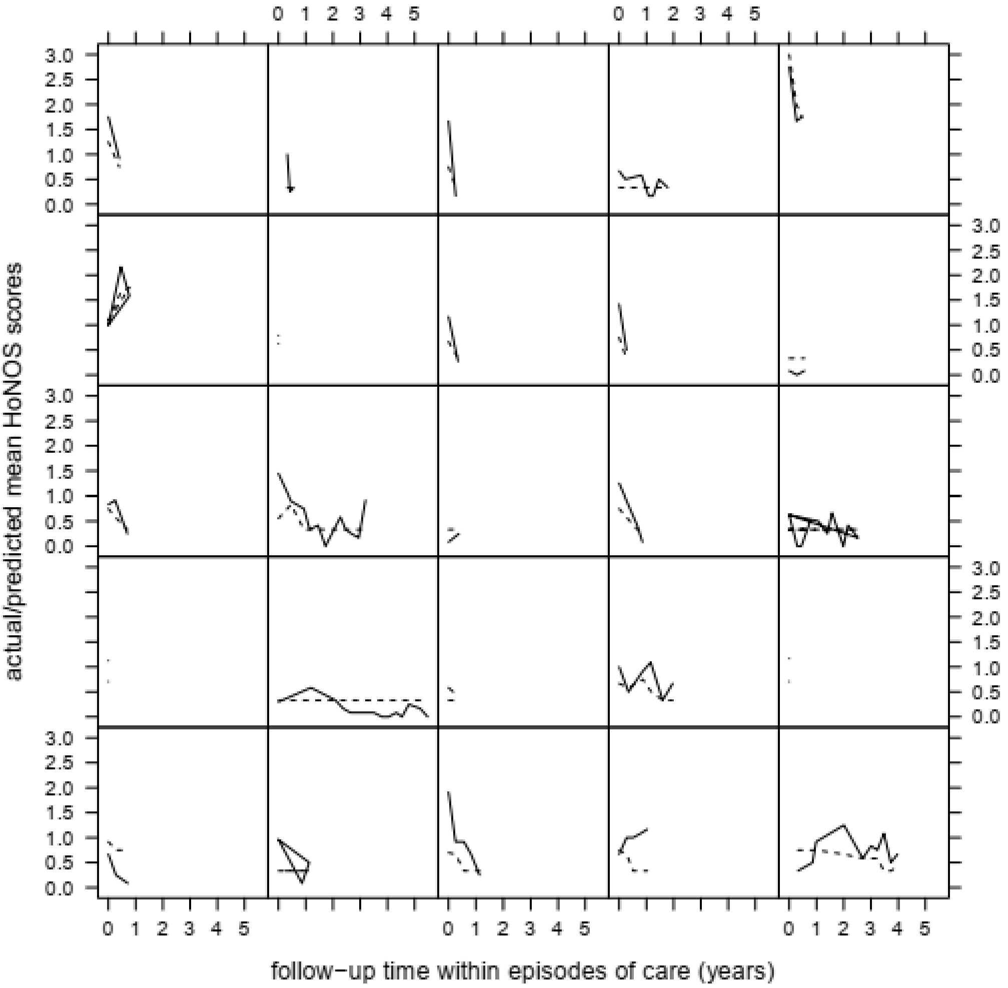


**Legend**: The figure shows the actual and posterior-predicted mean HoNOS scores for each of 25 individual patients (one patient in each panel). The predicted mean HoNOS scores derive from the Bayesian multi-level cumulative logit mixed model (mlclm). Note that three patients had two episodes of care and overlaps in their actual and predicted mean HoNOS scores from different episodes appear as triangles.

**Figure S4**: Posterior predictive check of adequate fit of the multi-level cumulative logit mixed model (mlclm)

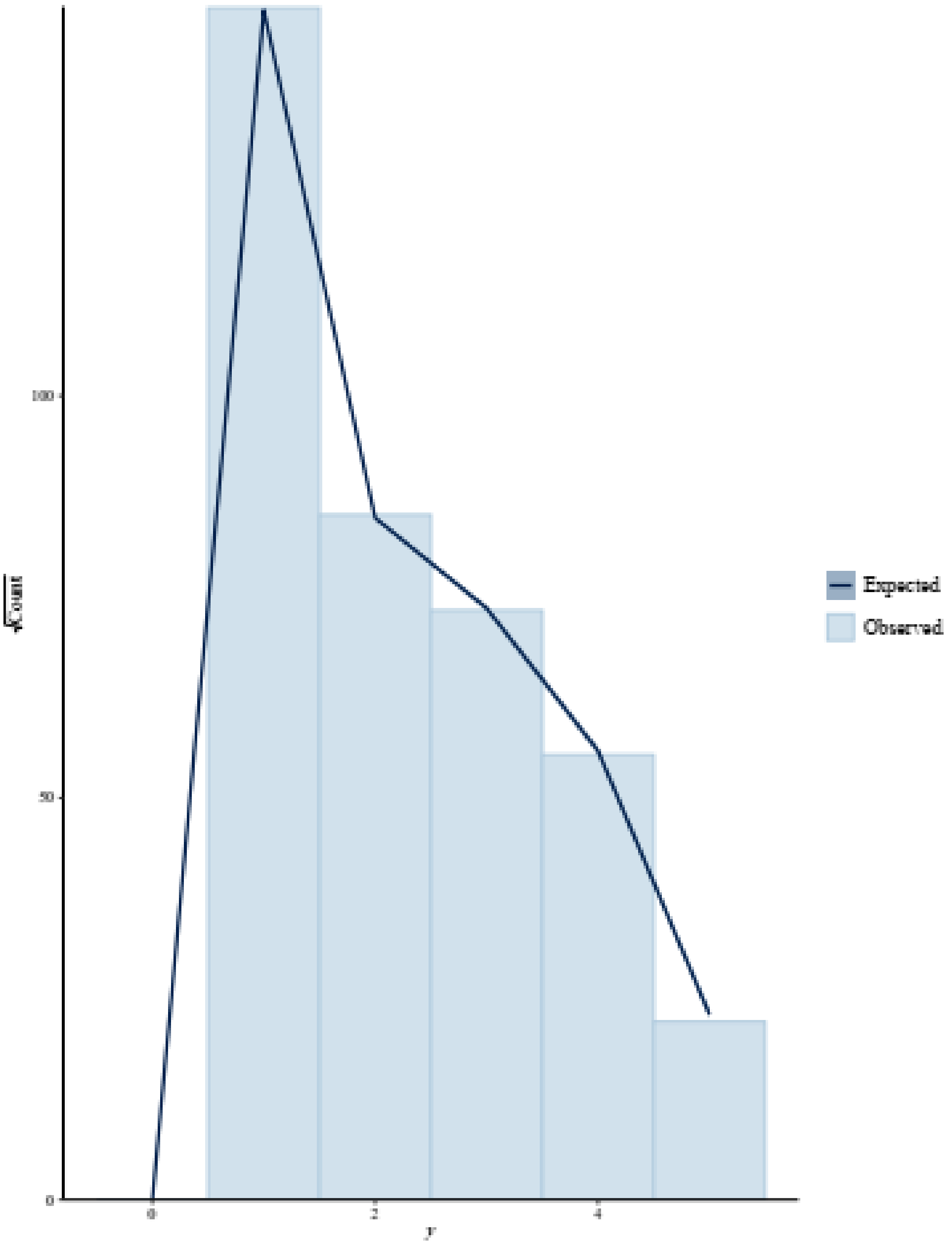


**Legend**: The figure shows the ‘rootogram’ for the mlclm, which compares the actual frequency of HoNOS ratings (bars along the x-axis, labelled ‘y’) with the mlclm’s predicted frequencies (line). Note that the scale of the y-axis shows the square root of the observed and expected counts of each rating.

**Figures S5a-c:** dependence of total HoNOS scores on estimates of sensitivity/bias of clinicians and severity of patients' problems from the multi-level cumulative link model (mlclm)

**Figure 5a**: dependence of overall mean HoNOS scores on clinician sensitivity/bias

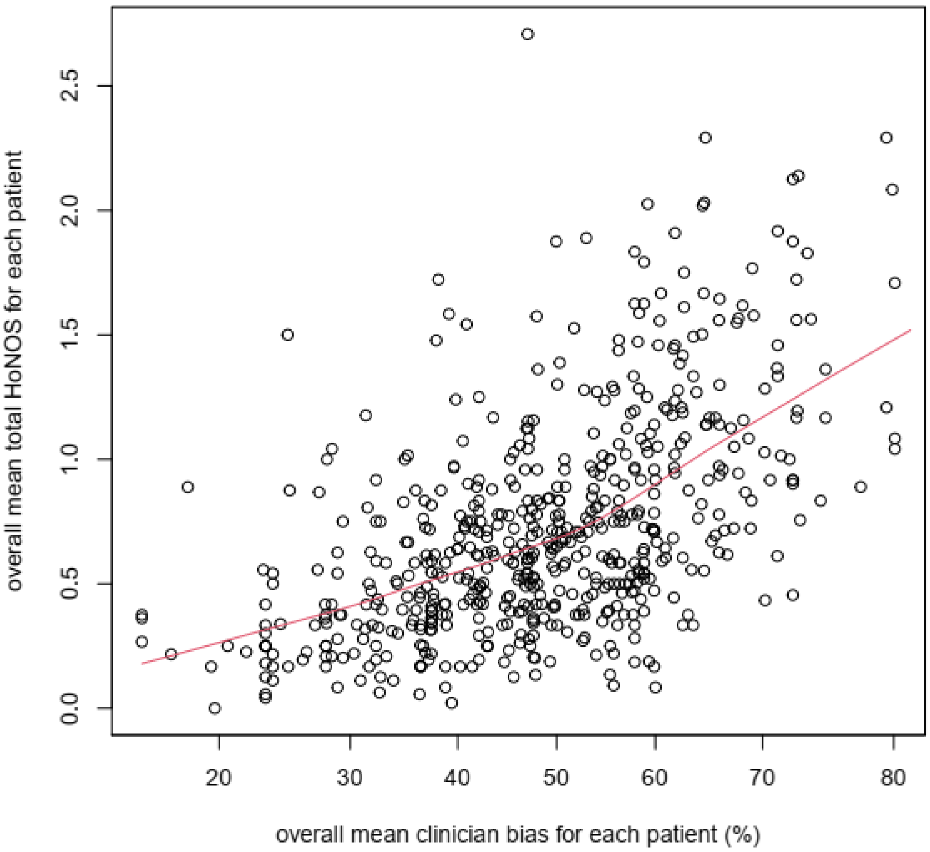


**Figure 5b**: dependence of mean HoNOS scores on initial severity of patients' problems

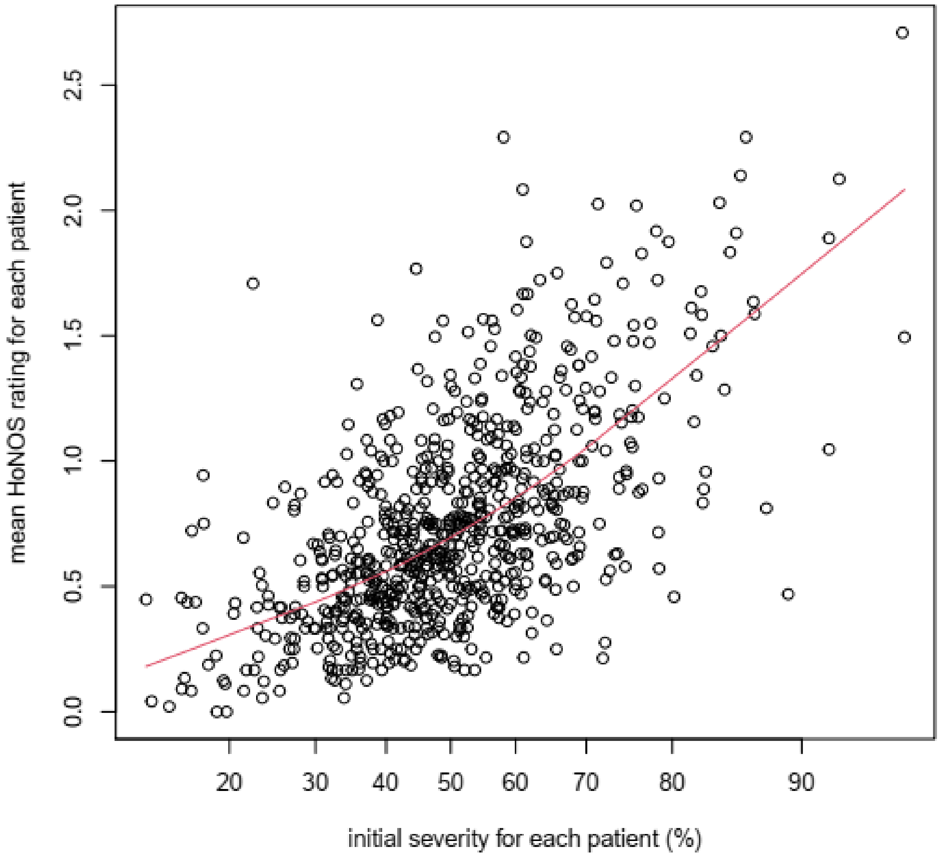

**Figure 5c:** dependence of mean HoNOS scores on rate of change of patients' problems

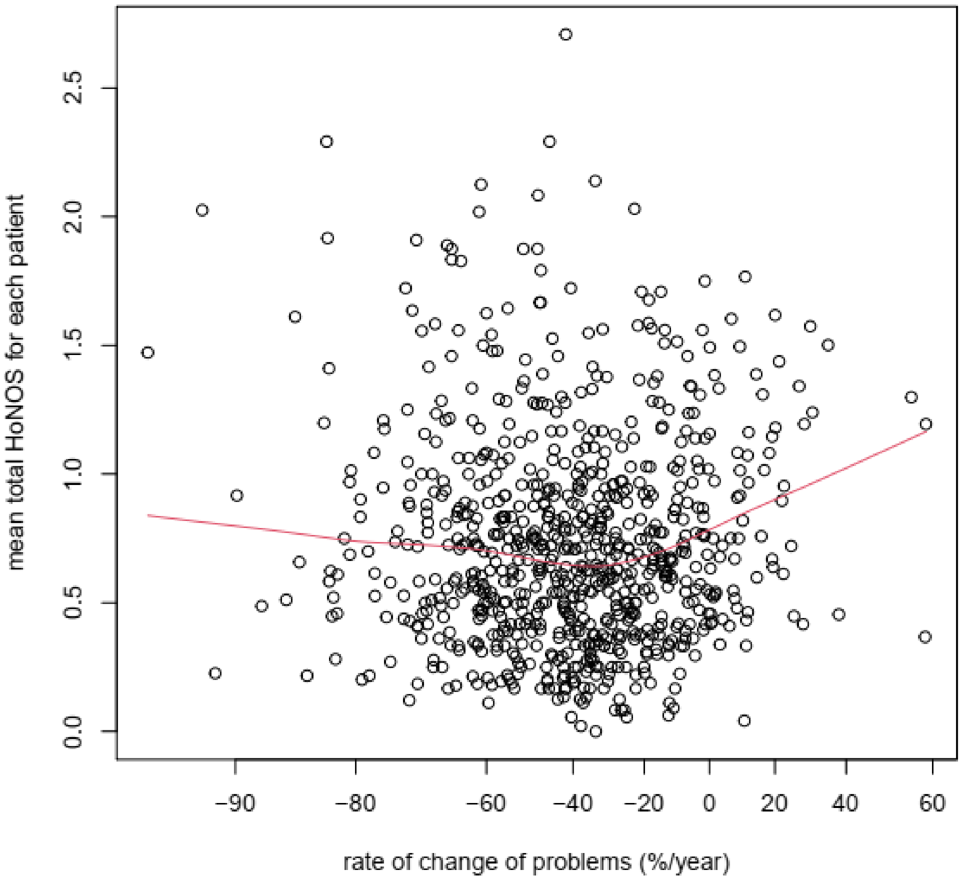


**Legend**: The y-axis in Figures S5a-c is the mean HoNOS rating. The x-axes for Figures S5a-c are (a) The mlclm's estimate of sensitivity/bias for each rater; (b) the mlclm's random intercept for each patient – which estimates the severity of each patient's problems at the initial assessment; (b) the mlclm's random slope over time for each patient – which estimates the rate of change of each patient's problems during follow-up within each episode of care (more negative values indicate faster resolution of problems). Values for the initial severity of patients' problems and clinician sensitivity are proportions (%), where the mlclm's mean is 50% (zero, on the logit scale). Values for the rate of change of patients problems are annual reductions of each patient's problems.

**Figures S6a-b:** orthogonality of estimates of the severity of patients' problems and of rater bias from the multi-level cumulative logit model (mlclm)

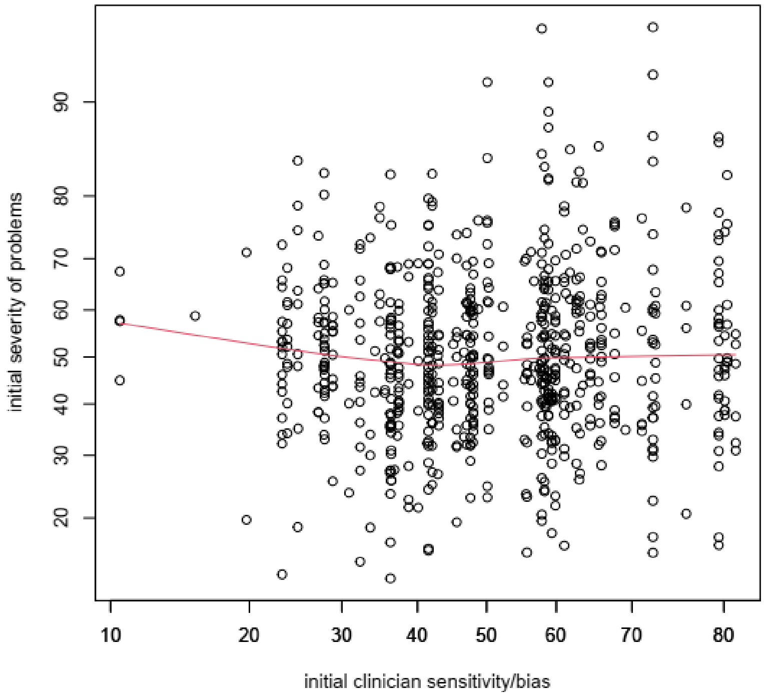


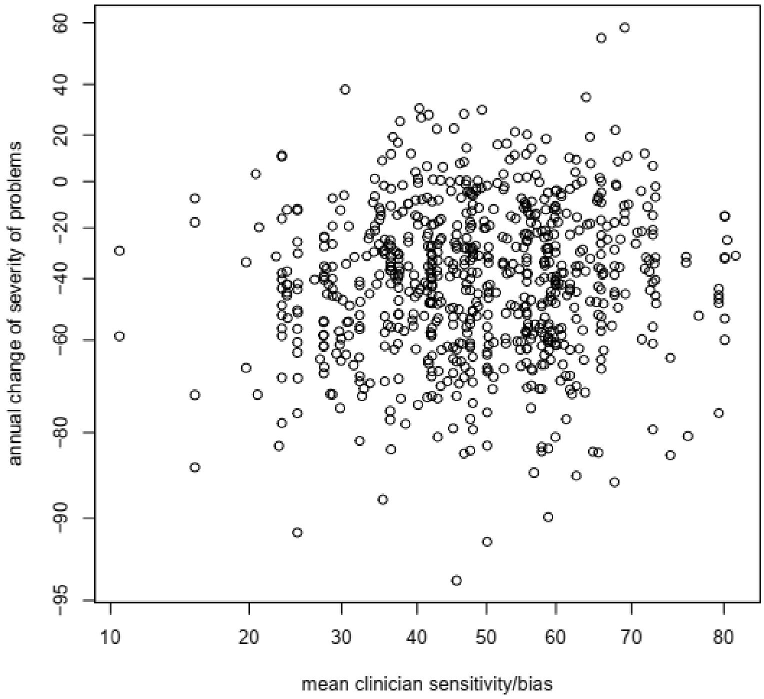


**Legend:** the x-axes in both figures show the sensitivity of the initial clinician who rated each patient. The y-axis in Figure S6a is the random intercept for each patient from the mlclm, which estimates the initial severity of each patient's problems. The y-axis in Figure S6b is the random slope over time for each patient, which estimates the rate of change of each patient's problems (further details as in legend of Figure S5).

**Figures 7a-b**: Caterpillar plots of the estimates of the severity of problems of individual patients

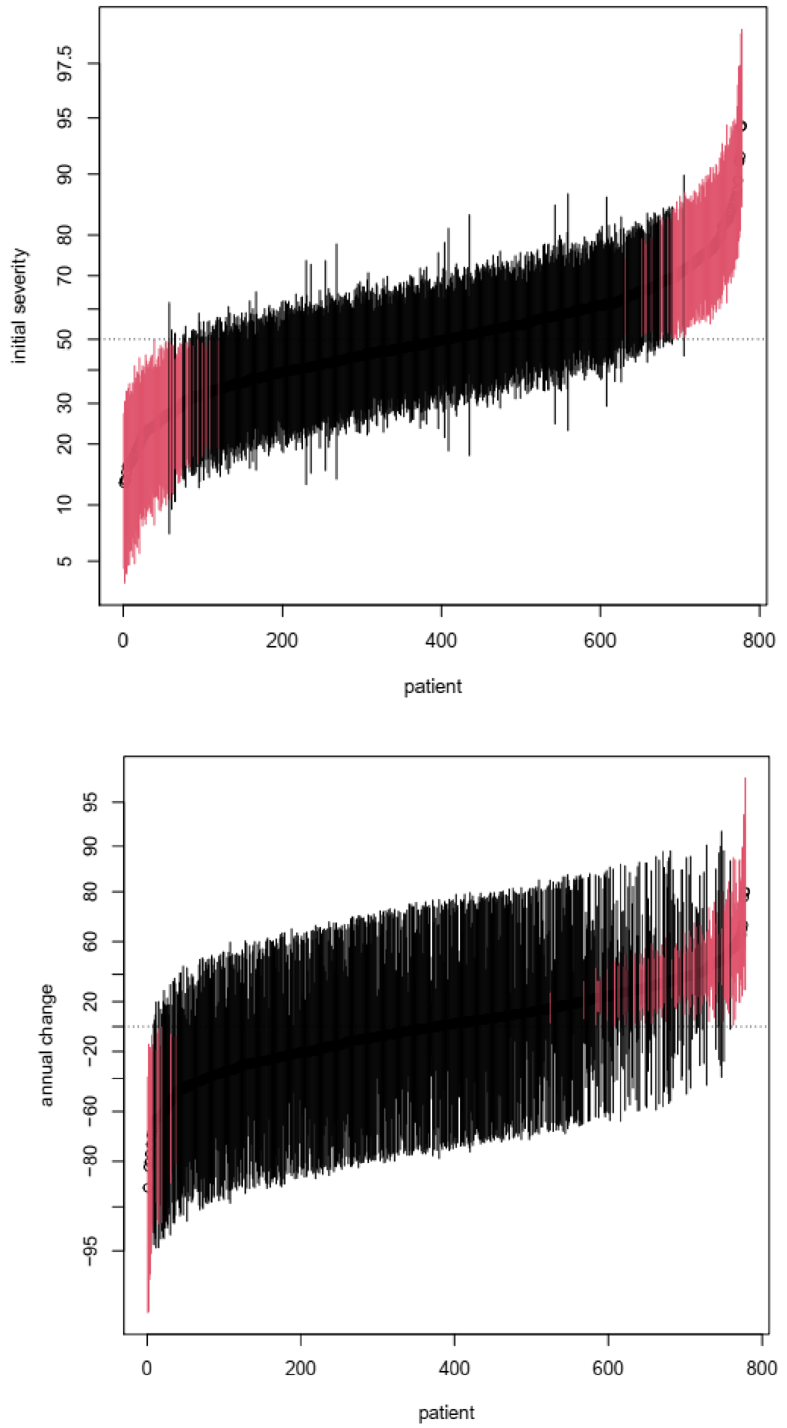


**Legend**: The x-axes show the index for each patient, ordered by increasing estimates of the severity of problems. CIs in red do not overlap the average value (defined as 50% = logit(0)). Figure 7a: The y-axis shows the estimate of initial severity of each patient's problems, together with its 95% Credible Interval from the multi-level cumulative link model (mlclm). Figure 7b: the y-axis shows the rate of change of each patient's problems (further details as in legend of Figure S5).

**Figures S8a-b**: boxplots of estimates of severity of patients' problems, grouped by CMHTs

**Figure 8a**: initial severity of patients' problems for each CMHT

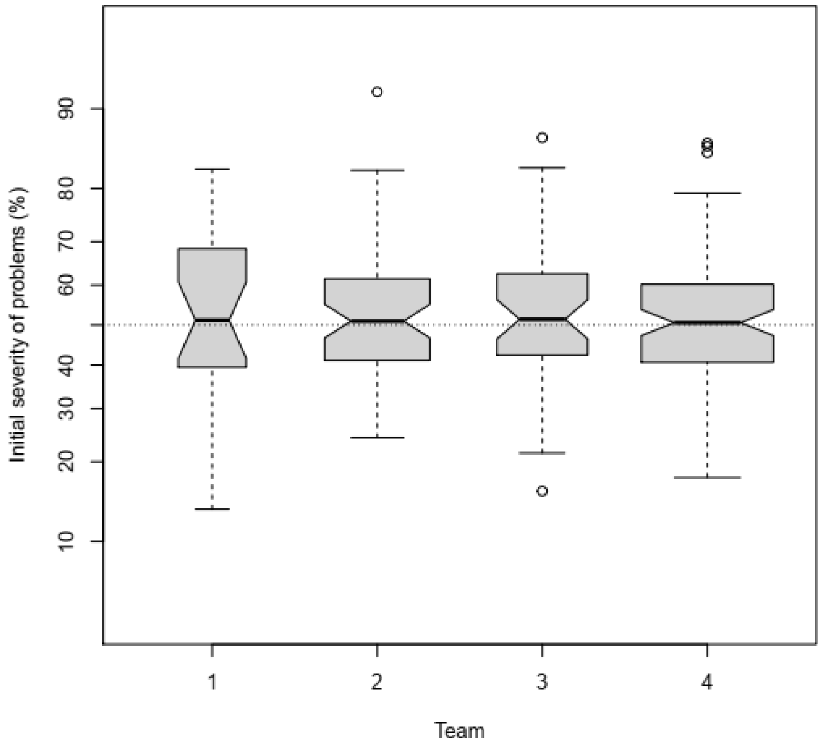


**Figure 8b**: annual change of patients' problems, for each CMHT

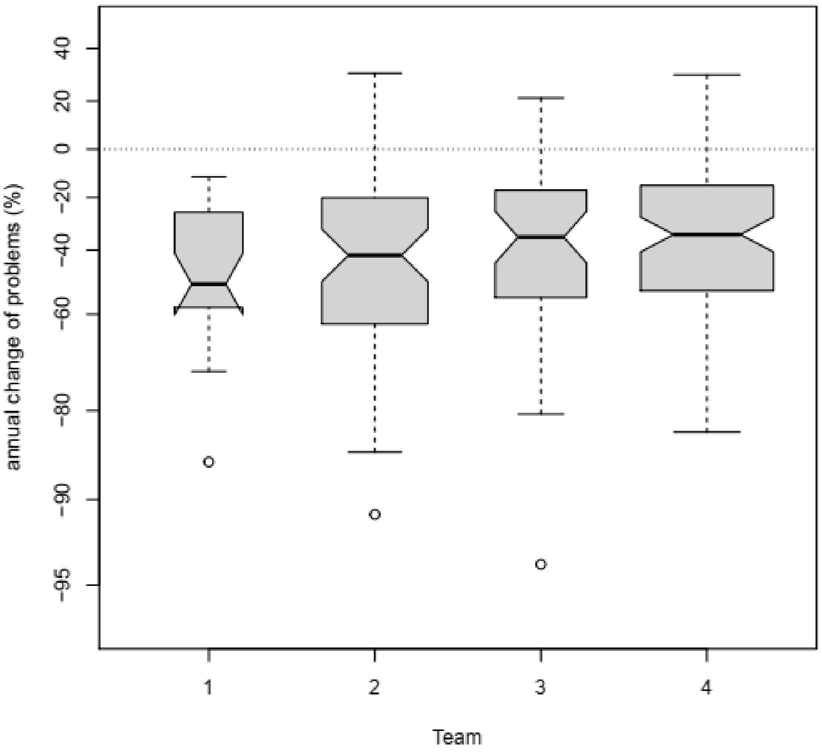


**Legend**: The notched box-and-whisker plots show the median and inter-quartile ranges of estimates of the severity of patients problems. The whiskers cover 99% of the range predicted from the inter-quartile range. The notches represent the confidence intervals for the medians.

**Figure S9**: Scatterplot of dependence of rate of change of patients' problems on their initial severity

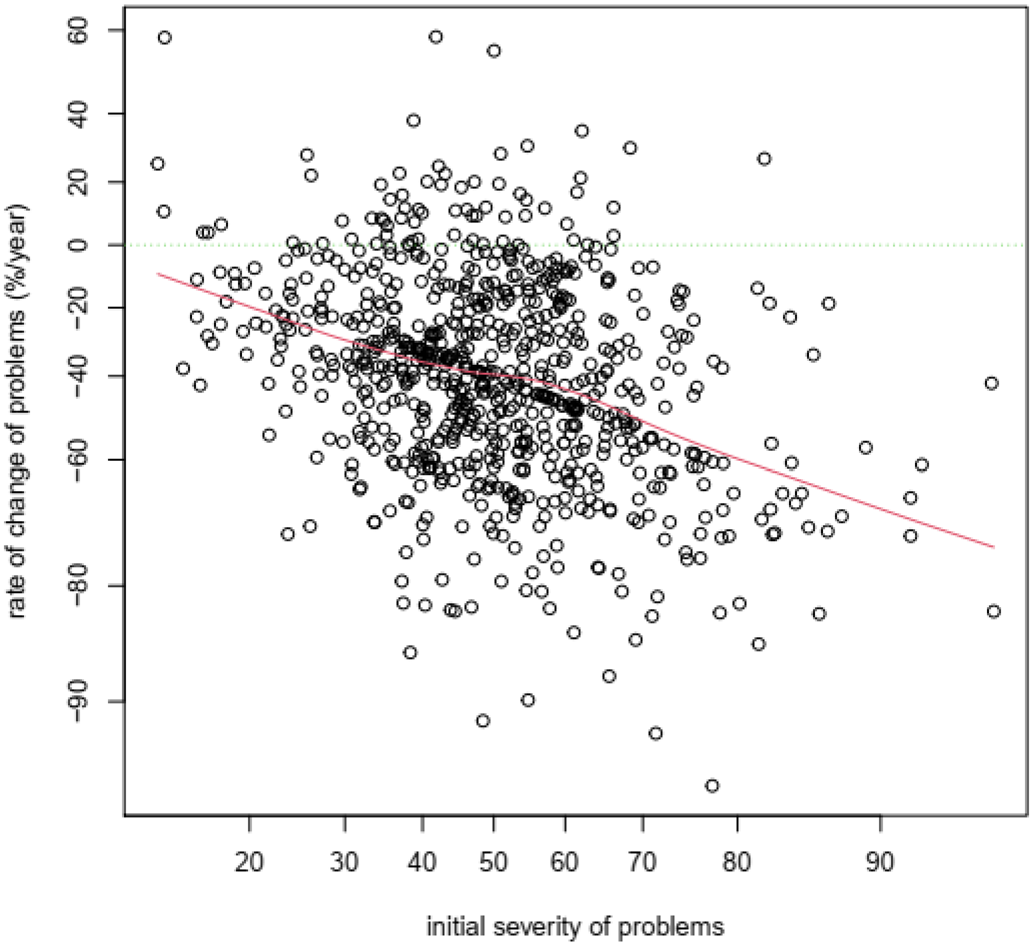


**Legend**: The y-axis is the rate of change of patients' problems (the mlclm's random slopes over time for individual patients). The x-axis is the initial severity of patients' problems (the mlclm's random intercepts for individual patients) (further details as in legend of Figure S5). The red line shows the non-parametric Lowess line that demonstrates the inverse relationship between the two outcomes.

**Figure S10**: Dependence of the hazard of inpatient admission on clinician sensitivity/bias

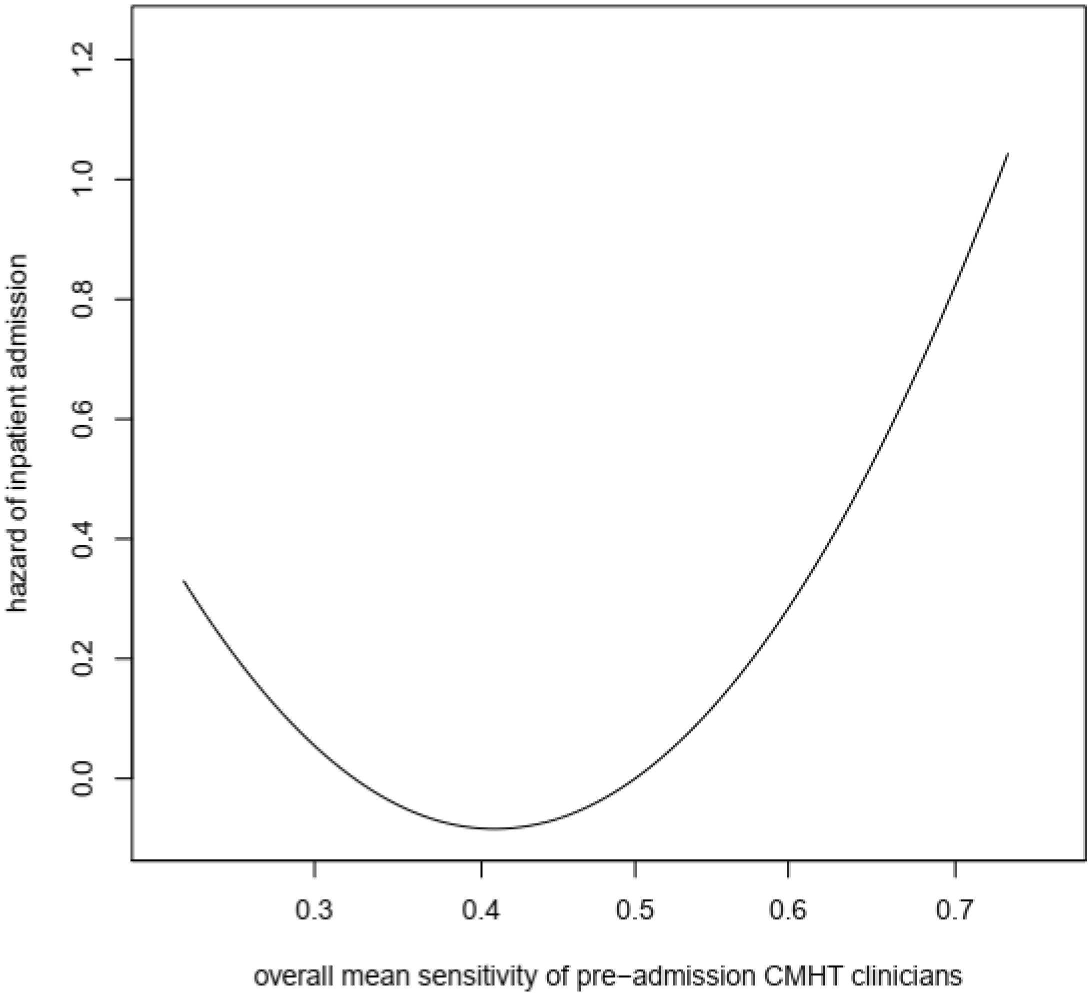


**Legend**: The y-axis shows the hazard of admission to the inpatient unit from the Cox's Proportional Hazards model. The x-axis shows the mean sensitivity of all pre-admission CMHT clinicians (see details in legend of Figure S5). The graph truncates the range of clinician sensitivities at 1% and 99% of the full observed range. Patients whose CMHT clinicians, overall, had the lowest or highest levels of sensitivity/bias had greater hazard of inpatient admission.

**SUPPLEMENTARY FILES**

note that <.rds> files require R for access

1) R code to generate the present results < R program file.txt > (2kB)

2) Multi-level cumulative link model (mlclm) – < NHL anon RN via NUTS 190920.rds > (255MB)

3) the data-set for the mlclm - < data for mlclm.txt > (3.1MB)

4) gamlss model of the dependence of rates of change of the severity of patients' problems on rater sensitivity and other variables – < ranptb ranrnsq anon 190920.rds > (1.7MB)

5) data set for the gamlss model - < gamlss data.txt > (87kB)

6) AFT model of the dependence of time to inpatient admission on rater sensitivity and other variables – AFT ranrnsq anon 190920.rds (8kB)

7) data-set for the AFT model - < AFT data .txt > (9kB)
